# Development of a Post-Acute Sequelae of COVID-19 (PASC) Symptom Lexicon Using Electronic Health Record Clinical Notes

**DOI:** 10.1101/2021.07.29.21261260

**Authors:** Liqin Wang, Dinah Foer, Erin MacPhaul, Ying-Chih Lo, David W. Bates, Li Zhou

## Abstract

**Objective:** To develop a comprehensive post-acute sequelae of COVID-19 (PASC) symptom lexicon from clinical notes to support PASC symptom identification and research.

**Methods:** We identified 26,117 COVID-19 positive patients from the Mass General Brigham’s electronic health records (EHR) and extracted 328,879 clinical notes from their post-acute infection period (day 51-110 from first positive COVID-19 test). The PASC symptom lexicon incorporated Unified Medical Language System^®^ (UMLS) Metathesaurus concepts and synonyms based on selected semantic types. The MTERMS natural language processing (NLP) tool was used to automatically extract symptoms from a development dataset. The lexicon was iteratively revised with manual chart review, keyword search, concept consolidation, and evaluation of NLP output. We assessed the comprehensiveness of the lexicon and the NLP performance using a validation dataset and reported the symptom prevalence across the entire corpus.

**Results:** The PASC symptom lexicon included 355 symptoms consolidated from 1,520 UMLS concepts. NLP achieved an averaged precision of 0.94 and an estimated recall of 0.84. Symptoms with the highest frequency included pain (43.1%), anxiety (25.8%), depression (24.0%), fatigue (23.4%), joint pain (21.0%), shortness of breath (20.8%), headache (20.0%), nausea and/or vomiting (19.9%), myalgia (19.0%), and gastroesophageal reflux (18.6%).

**Discussion and Conclusion:** PASC symptoms are diverse. A comprehensive PASC symptom lexicon can be derived using a data-driven, ontology-driven and NLP-assisted approach. By using unstructured data, this approach may improve identification and analysis of patient symptoms in the EHR, and inform prospective study design, preventative care strategies, and therapeutic interventions for patient care.

## BACKGROUND AND SIGNIFICANCE

As of July 2021, there had been over 190 million confirmed cases of the coronavirus disease 2019 (COVID-19) and four million deaths.^1^ The pandemic caused by the severe acute respiratory syndrome coronavirus 2 (SARS-CoV-2) is ongoing, and scientific understanding of its clinical stages and treatment strategies is evolving. Emerging prospective and retrospective studies suggest that some patients have persistent symptoms and/or develop delayed or long-term complications after their recovery from acute COVID-19, which may also be referred to as post-acute sequelae of SARS-CoV-2 infection (PASC) syndrome or long COVID.^2–4^ The onset, scope, and duration of PASC symptoms that often involve multiple organ systems represent a new phase of the pandemic, with significant implications for health care delivery.^5^ Efficient, scalable tools to identify and analyze patient PASC symptoms are essential to inform risk assessment, prevention and treatment strategy development, and outcome estimation.^6, 7^

The characterization of PASC symptoms has varied widely by study.^8^ This heterogeneity of research findings is attributed to multiple factors, including variation in study design, patient populations, the “post-acute COVID-19” timeframe, and sample size.^2, 8–15^ Most early studies on PASC symptoms relied on patient survey data, manual chart review, and in person follow-up.^6, 12, 13, 15–18^ These studies were often limited by sample size and reporting biases. Longitudinal electronic health record (EHR) data serve as a rich data source for studying PASC symptoms. However, retrospective studies that primarily utilize structured EHR data (e.g., lab results or International Classification of Disease [ICD] diagnosis codes)^11, 14, 19^ may miss many clinical symptoms, which are often documented in clinical notes.

Natural language processing (NLP) can automatically identify relevant symptoms and complications at different clinical stages from large volumes of longitudinal notes of a large patient cohort.^20, 21^ Several NLP approaches have been developed to extract COVID-19 signs or symptoms using either lexicon-based or machine learning-based approaches.^22, 23^ However, these approaches have focused on acute phases of COVID-19, while post-acute and long-term symptom identification were not covered and remain an unmet need. A significant challenge for developing such an NLP tool is the wide variation in potential post-acute COVID-19 symptoms, as COVID-19 survivors can experience a heterogeneous constellation of respiratory, cardiovascular, neurologic, psychiatric, dermatologic, and gastrointestinal symptoms and/or complications. A comprehensive lexicon encoded with a standard medical terminology that encompasses a broad range of PASC symptoms derived from large volumes of EHR notes is crucial for NLP tool development and utility and future EHR-based PASC analytics and research.

### OBJECTIVE

Our objective of this study was to develop a comprehensive PASC symptom lexicon using medical ontology-based and data-driven approaches. We used a large biomedical thesaurus, a large clinical EHR dataset of COVID-19 cases, an NLP system, and iterative manual chart review to develop, improve, and evaluate the lexicon.

## MATERIALS AND METHODS

We used a two-phase approach to develop and evaluate a post-acute COVID-19 symptom lexicon (**Figure 1**).

**Figure 1.**
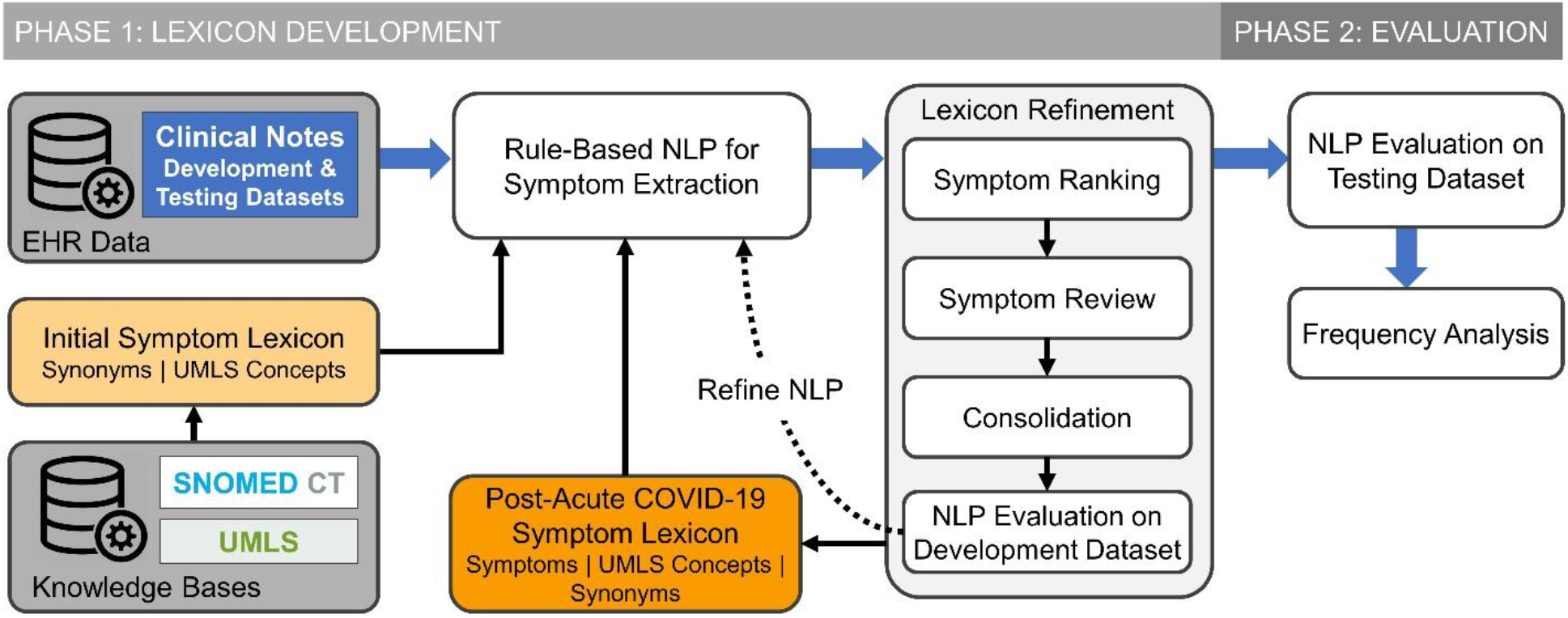
Schematic diagram of PASC symptom lexicon development and evaluation in a natural language processing (NLP) system.

### Settings and Data Sources

The study was conducted in the Mass General Brigham (MGB) healthcare system, the largest integrated healthcare delivery system in New England. It includes academic tertiary care medical centers, community hospitals, outpatient primary care, and specialty care offices. MGB maintains an enterprise data warehouse which is a centralized repository of EHR data, including demographics, encounters, diagnoses, problem lists, laboratory results, medications, flowsheets, and clinical notes (e.g., inpatient and outpatient encounters, discharge summaries, telephone calls, patient electronic messages). This study was approved by the institutional review board of MGB with waiver of informed consent from study participants for secondary use of EHR data.

### Study Cohort and Data Extraction

We identified patients who were ≥18 years of age and had a positive test result for SARS-CoV-2 by polymerase chain reaction (PCR) clinical assay between March 4, 2020 and February 09, 2021. We defined days 0-40 from first positive PCR test as the acute COVID-19 phase,^24^ days 41-50 as a “grace” period to allow for additional results from testing conducted during the acute period, and day 51 and after as the post-acute COVID-19 period. We used all clinical notes from days 51 to 110 (a total of 60 days) during the post-acute period for each patient.^13, 25, 26^ Patients who died before day 51 were excluded from the study. We divided the corpus of clinical notes into development (90% of the study cohort) and validation (10% of the study cohort) datasets to meet our study objectives.

### Phase 1: PASC Symptom Lexicon Development

In the first phase of this study, using the Unified Medical Language System^®^ (UMLS) and SNOMED CT, we first compiled an initial symptom lexicon containing selected UMLS concepts and related synonyms. Next, we applied a rule-based NLP algorithm to extract mentions of the symptoms in the lexicon from the development dataset. We refined the lexicon based on an iterative process including: (1) symptom ranking based on prevalence; (2) manual review to identify missed post-acute COVID-19 symptoms; (3) consolidation of concepts of similar meaning; (4) evaluation of NLP performance.

#### Initial Symptom Lexicon Development

We used a knowledge-based approach to develop an initial lexicon inclusively containing symptom concepts and their synonyms that may appear in clinical notes. First, we compiled an initial list of concepts from UMLS Metathesaurus^®^ (version 2020AB) and SNOMED CT Core Problem List Subset (version February 2021).^27^ We included concepts from UMLS Metathesaurus in the English language, with vocabulary source of “SNOMEDCT_US”, and with semantic types under any of the following categories: “Pathologic Function”, “Finding”, and “Anatomical Abnormality”.^28^ We also included concepts from the SNOMED CT Core Problem List Subset with indication of the concepts as “finding” or “disorder.” From the resulting concept list, we extracted synonyms from the UMLS Metathesaurus as part of the initial lexicon.

#### Symptom Extraction

For post-acute COVID-19 symptom extraction, we adapted the Medical Text Extraction, Reasoning, and Mapping System (MTERMS),^29^ a multipurpose NLP tool, containing modules such as a section identifier, a sentence splitter, a lexicon-based search module, and a rule-based concept extractor. We used MTERMS’s section identifier to process clinical notes in the development dataset, excluding sections less likely to contain patients’ post-acute COVID-19 symptoms, such as medications list, laboratory results, immunization history, hospital discharge instructions, plan of care, and family history. Next, we split the sections into sentences and applied MTERMS’s lexicon-based search module. This search module can utilize a pre-defined lexicon to match mentions of terms contained in the lexicon in the text. Synonyms in the symptom lexicon were considered as NLP search terms, and each mention of the terms was mapped to a UMLS concept. Those terms were further processed by a series of manually constructed or adapted rules to exclude invalid symptoms. For example, the MTERMS negation module, which adopted rules from Negex,^30^ was further applied to exclude negated terms (e.g., “no fever”). We also used MTERMS’s modifier module to identify experiencers other than the patient (e.g., mother, father, wife, husband, son, daughter). We used an allergen lexicon to identify and exclude symptoms mentioned in the context of drug-induced adverse events, such as drug allergic reactions (e.g., ‘ACE Inhibitors Angioedema’, ‘Metformin GI Upset’).^31^ Finally, we used a pattern-matching approach to identify sentences in which symptoms were mentioned as part of risk factors, side effects, instructions, and survey questions. The NLP rules were iteratively added or refined during the lexicon development stage to optimize NLP performance.

#### PASC Symptom Ranking and Review

PASC symptoms were defined as a patient feature occurring in the post-acute COVID-19 period. We distinguished patient symptoms from objective findings (e.g., laboratory results, blood pressure measurements) and provider-based disease diagnoses (e.g., choledocolithiasis). This enabled inclusion of symptoms such as palpitations, but not the corresponding diagnoses of “Wolff-Parkinson-White syndrome” which require provider-based assessment, criteria, and evaluation. At each stage of development, we used the current symptom lexicon to extract all possible symptoms in clinical notes. We ranked all the concepts by frequency of patient occurrence. We manually reviewed the concepts to identify symptom-specific terms, as defined above. For example, to identify symptoms related to “problem with smell or taste”, we manually reviewed clinical notes for mentions of “smell” and “taste” and added additional 143 synonyms to the lexicon. Given the long tail of symptoms distribution, we prioritized identification of the most common symptoms in the patient cohort. Concepts occurring in 50 or more patients were manually reviewed by two clinicians (DF and YL). Concepts occurring in less than 50 patients were manually searched using the symptom keywords (identified from the set of concepts occurring in more than 50 patients), e.g., “pain”, “swelling”. This enabled us to maximize symptom capture and facilitate subsequent concept consolidation even for rarer symptoms. Conflicts in classification were resolved by consensus review; final determination was made by LW.

#### Symptom Consolidation

Among the identified symptom concepts, we consolidated those with clinically similar meanings into groups. This minimizes clinically insignificant duplication in the symptom term list and improves the analysis of symptom patterns and trends. For example, ‘paresthesia’ (CUI: C0030554) and ‘pins and needles’ (CUI: C0423572) are included in the UMLS Metathesaurus as two different concepts; however, from a symptom-oriented perspective “pins and needles” is a manifestation of paresthesia and was consolidated under “Paresthesia”. In clinical notes, some concepts were frequently paired in a single term, such as “loss of smell and taste” or “nausea and/or vomiting.” To prevent symptom duplication across multiple unique terms (e.g., “nausea” “vomiting” “nausea and vomiting” “nausea or vomiting” “nausea and/or vomiting”) we refined the lexicon to reflect the clinical context and merged the terms into a single concept, “nausea and/or vomiting”. For some general symptoms, we consolidated the concepts based on anatomic site. For example, pain is a common symptom in the development dataset with over 300 concepts containing the word “pain”. We manually consolidated the concepts by pain site, resulting in several concept groups, e.g. “abdominal pain”, “joint pain”, “pain in extremities”, “chest pain”.

#### NLP Evaluation and Lexicon Refinement

Based on the steps described above we were able to iteratively refine the symptom lexicon, further enhanced by the final step of NLP evaluation. With each lexicon revision, we evaluated the performance of NLP for its ability to identify symptoms from a random set of clinical notes in the development dataset. Through this evaluation, the identified false positive (e.g. missed negation) and false negative symptoms enabled us to refine the symptom lexicon and NLP rules. For example, based on the UMLS Metathesaurus, the term ‘fit’ is a colloquial synonym for seizure, and ‘TEN’ is an acronym for ‘toxic epidermal necrolysis’. However, without advanced word sense disambiguation functions, these terms could cause inaccurate extraction or false positives. Therefore, term mapping rules were refined to minimize false positive symptom identification. Additional examples of challenges in lexicon development are described in **Supplementary eTable 1**.

### Phase 2: Evaluation of Symptom Lexicon

Once the lexicon reached a satisfactory level of performance during the NLP evaluation in the development dataset, we moved on to the second phase, which evaluated its performance in the NLP system using the validation dataset of a different cohort to gain insights about the lexicon.

#### Assessing NLP Performance in Identifying PASC Symptoms

With the final lexicon, we evaluated NLP performance in identifying post-acute COVID-19 symptoms from clinical notes in the validation dataset. We measured performance in terms of precision (or positive predictive value) for the 50 most common symptoms and recall (or sensitivity) across all the symptoms. To calculate precision, we randomly selected 50 sentences for each symptom from the validation dataset (a total of 2,500 sentences) and manually identified the number of true positive symptoms. To calculate recall across all symptoms in the lexicon, we randomly selected 50 clinical notes from the validation dataset and manually identified symptom terms. The notes were also processed by the NLP tool for symptom extraction. We then counted the number of manually annotated symptoms (N) and the number of symptoms extracted by both manual (N) and NLP (n) review and calculated the ratio of n and N for recall.

#### Frequency Analysis

After validation, we applied the lexicon and NLP to the entire clinical note corpus. We calculated the frequency of PASC symptoms within the entire study population during the 2-month follow-up.

## RESULTS

Overall, 51,485 adult patients met initial inclusion criteria for the study, among which, 26,117 (50.7%) had 328,879 clinical notes during the follow-up period. In total, 23,505 (90%) patients representing 299,140 notes were included in the development dataset and 2,612 (10%) patients with 29,739 notes were included in the validation dataset. Basic demographic characteristics are summarized in **Table 1**.

**Table 1.** Demographic characteristics of patients in the development and validation datasets

| **Characteristic** | **Patient Cohort**<br>**n=26,117** | **Patients in development dataset**<br>**n=23,505** | **Patients in validation dataset**<br>**n=2,612** |
| --- | --- | --- | --- |
| Age, Mean (SD), y | 51.6 (18.2) | 51.6 (18.2) | 51.5 (18.4) |
| Female, No. (%) | 16177 (61.9) | 14,578 (62.0) | 1,599 (61.2) |
| Race* |  |  |  |
| White | 17,752 (68.0) | 15,939 (67.8) | 1,813 (69.4) |
| Black | 2,551 (9.8) | 2,289 (9.7) | 262 (10.0) |
| Asian | 704 (2.7) | 630 (2.7) | 74 (2.8) |
| Other/unknown | 5110 (19.6) | 4647 (19.8) | 463 (17.7) |
| Ethnicity, Hispanic* | 5632 (21.6) | 5,105 (21.7) | 527 (20.2) |
| Clinical Notes, count | 328,879 | 299,140 | 29,739 |

\* Self-reported.

The initial lexicon based on the selected UMLS sematic types included a total of 157,245 unique concepts and 604,056 synonyms. Eleven percent (n=17,701) of these concepts were mentioned in the development dataset. Manual review of the 2,660 concepts that occurred in 50 or more patients during the study period identified 698 (26.1%) concepts as symptom related. Review of the 15,041 UMLS concepts occurring in less than 50 patients identified an additional 822 symptom concepts. Consolidation among the total 1,520 symptom concepts resulted in a final count of 355 symptoms. **Table 2** displays symptoms from the lexicon, corresponding consolidated UMLS concepts, and examples of symptom synonyms from clinical notes. The complete symptom lexicon is available on GitHub.^32^

**Table 2.** Selected examples of post-acute COVID-19 symptoms, consolidated Unified Medical Language System (UMLS) concepts, and synonyms from electronic health record clinical notes

| Symptoms | Consolidated UMLS concepts | Examples of Synonyms in Clinical Notes |
| --- | --- | --- |
| Fatigue | C0015672:Fatigue, C0231218:Malaise, C0015674:Chronic Fatigue Syndrome, C0023380:Lethargy, C0392674:Exhaustion, C0024528:Malaise And Fatigue, C0424585:Tires Quickly, C0849970:Tired, C0439055:Tired All The Time, C2732413:Postexertional Fatigue, C3875100:Fatigue Due To Treatment, C4075947:Occasionally Tired | Fatigue, tiredness, malaise, tired, fatigued, lethargy, ill feeling, feeling unwell, feel tired, lethargic |
| Loss of appetite | C0003125:Anorexia Nervosa, C0232462:Decrease In Appetite, C0426587:Altered Appetite, C1971624:Loss Of Appetite, C0566582:Appetite Problem | Loss of appetite, decreased appetite, poor appetite, appetite changes, change in appetite, decrease in appetite, appetite loss |
| Sleep apnea | C0037315:Sleep Apnea, C0003578:Apnea, C0020530:Hypersomnia With Sleep Apnea, C0751762:Primary Central Sleep Apnea, C1561861:Organic Sleep Apnea, C2732337:Sleep Hypoventilation | Sleep apnea, apneas, apnea, sleep disturbance, sleep disturbances, sleep problems, sleep disorder |
| Sinonasal congestion | C0027424:Nasal Congestion, C0152029:Congestion Of Nasal Sinus, C0240577:Swollen Nose, C0700148:Congestion, C0439030:C/O Nasal Congestion, C0522564:Chronic Congestion | Congestion, nasal congestion, sinus congestion, stuffy nose, congested nose |
| Problem with smell or taste | C0003126:Loss Of Sense Of Smell, C0013378:Taste Sense Altered, C0039338:Disorder Of Taste, C0234259:Sensitive To Smells, C0240327:Metallic Taste, C0423564:Abnormal Taste In Mouth, C0423570:Unusual Smell In Nose, C0481703:Problem With Smell Or Taste, C0553757:Disorder Of Smell, C0578994:Unpleasant Taste In Mouth, C1510410:Sense Of Smell Altered, C2364082:Sense Of Smell Impaired, C2364111:Loss Of Taste | Loss of smell, anosmia, loss of smell or taste, loss of smell / taste, loss of taste or smell, loss of taste and smell, loss of taste, dysgeusia, loss of smell and taste, loss of sense of taste or smell, metallic taste, taste changes, loss of sense of smell, decreased sense of smell, loss of taste / smell, loss of sense of smell or taste, taste loss, ageusia, smell changes, change in sense of smell or taste, hyposmia, loss of sense of taste / smell |

**Table 3** shows the 50 most common symptoms in the entire patient cohort and their prevalence in clinical notes. Symptoms with the highest frequency included pain (43.1%), anxiety (25.8%), depression (24.0%), fatigue (23.4%), joint pain (21.0%), shortness of breath (20.8%), headache (20.0%), nausea and/or vomiting (19.9%), myalgia (19.0%), and gastroesophageal reflux (18.6%). Using the final PASC symptom lexicon, the NLP performance in clinical note symptom extraction for individual symptoms was measured in the validation dataset. 46 concepts (92%) had precision measured above 0.90; average precision was 0.94 (range, 0.82 to 1.0). To calculate recall, a total of 1,481 sentences were reviewed from 50 notes. Manual review identified 104 symptom terms, among which NLP identified 87 symptoms. Therefore, the estimated recall of the final PASC symptom lexicon in our NLP system was 0.84. Our error analysis revealed that the false negative cases were caused by the following reasons: missing abbreviations (e.g., ‘OSA’ for obstructive sleep apnea, ‘HA’ for headache), uncommon synonyms (‘short tempered’), and misspellings (e.g., ‘pian’ for pain).

**Table 3.** 50 most common post-acute COVID-19 patient symptoms in electronic health record clinical notes by symptom frequency, and corresponding precision of natural language processing (NLP) performance for unique symptom extraction

| **Top 1-25 Symptoms** | **% frequency of symptoms** | **Precision** | **Top 26-50 Symptoms** | **% frequency of symptoms** | **Precision** |
| --- | --- | --- | --- | --- | --- |
| Pain | 43.1 | 0.94 | Insomnia | 11.2 | 0.94 |
| Anxiety | 25.8 | 0.98 | Pain in extremities | 10.7 | 1.0 |
| Depression | 24.0 | 0.90 | Paresthesia | 10.7 | 0.92 |
| Fatigue | 23.4 | 1.0 | Peripheral edema | 10.5 | 0.98 |
| Joint pain | 21.0 | 0.98 | Palpitations | 10.3 | 0.94 |
| Shortness of breath | 20.8 | 0.94 | Diarrhea | 10.3 | 0.92 |
| Headache | 20.0 | 0.92 | Itching | 9.4 | 0.92 |
| Nausea and/or vomiting | 19.9 | 1.0 | Erythema | 9.2 | 0.98 |
| Myalgia | 19.0 | 0.96 | Lower urinary tract symptoms | 8.7 | 0.98 |
| Gastroesophageal reflux | 18.6 | 0.94 | Lymphadenopathy | 8.3 | 0.96 |
| Cough | 17.5 | 0.92 | Edema | 7.9 | 0.88 |
| Back pain | 16.9 | 0.98 | Weight gain | 7.3 | 0.98 |
| Stress | 15.1 | 0.86 | Sinonasal congestion | 7.1 | 0.96 |
| Fever | 14.7 | 0.94 | Pain in throat | 6.4 | 0.98 |
| Swelling | 14.7 | 0.90 | Abnormal gait | 5.9 | 1.0 |
| Bleeding | 14.7 | 0.90 | Respiratory distress | 5.8 | 0.82 |
| Weight loss* | 14.2 | 0.98 | Visual changes | 5.8 | 0.92 |
| Abdominal pain | 14.1 | 0.98 | Chills | 5.6 | 0.86 |
| Dizziness or vertigo | 14.0 | 0.94 | Urinary incontinence | 5.6 | 0.96 |
| Chest pain | 12.5 | 0.90 | Sleep apnea | 5.4 | 0.94 |
| Weakness | 12.3 | 0.94 | Confusion | 5.4 | 0.98 |
| Constipation | 11.9 | 0.96 | Hearing loss | 5.2 | 1.0 |
| Skin lesion | 11.9 | 0.94 | Problem with smell or taste | 5.0 | 0.94 |
| Wheezing | 11.9 | 0.98 | Difficulty swallowing | 4.9 | 0.98 |
| Rash | 11.4 | 0.82 | Loss of appetite | 4.8 | 0.96 |

## DISCUSSION

We developed a comprehensive lexicon of post-acute COVID-19 symptoms from the EHR and validated the NLP tool using the lexicon for symptom extraction; the lexicon is publicly available. This work advances the study of post-acute COVID-19 symptoms by providing a systematic approach and scalable tool to identify patient symptoms post-COVID-19 infection in large datasets. Free-text clinical notes represent an underutilized data source in the active field of post-acute COVID-19 research; this study will facilitate knowledge-based NLP approaches to identifying post-COVID-19 symptoms. Subsequently, these symptoms can be used to characterize patient populations or organ/system-based domains for targeted preventative or therapeutic interventions, and support future research.

Our study has several strengths. We used clinical notes to study post-acute COVID-19 symptoms. This is distinct from prior studies that have used other EHR-based data elements such as billing/diagnosis codes, laboratory results, or medications.^14, 19, 33^ Billing/diagnosis codes may represent disease states encompassing a variety of symptoms, and therefore may underestimate each unique symptom and trends across symptoms. Clinical notes also have an advantage over these sources for studying symptoms, by representing the clinical encounter between patient and provider, and capturing patient reported symptoms--including outside of the formal visit setting (e.g., patient messages and telephone encounters). While patient surveys also capture symptoms, they are subject to responder bias and limited in scale.^2^ In larger survey studies across institutions or from social media-based sources, acute COVID-19 infection status may be difficult to ascertain.^34^

Our study also used an NLP-based approach which is well-suited to systematically process unstructured data containing post-acute COVID-19 symptoms.^8^ Compared to machine learning-based approaches for symptom extraction,^23^ which may focus on specific symptoms as unique classification tasks, requiring resource-intensive document annotation, a knowledge-based NLP approach can leverage existing knowledge bases to investigate and identify a wide variety of symptoms from a large dataset. This has also made our study distinct from previous work studying post-COVID-19 sequalae in specific patient populations (e.g., with neuropsychiatric outcomes)^14^ or following a specific level of COVID-19 infection acuity (e.g., necessitating ICU admission).^10^ Our study used data from a general population for lexicon development.^33^ This supports comprehensive identification of symptoms for the lexicon across a medically diverse population; future studies may then compare symptom prevalence and frequency in distinct populations.

Notably the most common symptoms identified by our NLP-based approach have significant overlap with previously identified post-acute COVID-19 findings in meta-analysis studies of surveys and observational data.^25^ Lopez-Leon et al. conducted a systematic literature review and identified more than 50 long-term effects of COVID-19,^2^ most commonly including fatigue, headache, attention disorder, hair loss, and dyspnea. Halpin et al. identified fatigue, breathlessness, anxiety/depression, concentration problems, and pain among the five most common post-discharge symptoms in 100 patients hospitalized with COVID-19 (ward and ICU).^16^ A recent analysis of new ICD-coded outpatient diagnoses (including symptom-related codes) among non-hospitalized COVID-19 patients 28-108 days post COVID-19 diagnosis listed pain in throat and chest, shortness of breath/dyspnea, headache, malaise, and fatigue as the top five symptoms determined to be potentially related to COVID-19.^19^ Our study identified the most common 10 symptoms as pain, anxiety, depression, fatigue, shortness of breath, joint pain, nausea and/or vomiting, headache, myalgia, and gastroesophageal reflux, highlighting symptoms common to both inpatient and outpatient-based studies and raising additional symptoms for consideration. This likely reflects our inclusion of a broad post-acute COVID-19 population and a focus on specific symptoms, rather than “findings” or “effects” which may capture disease states that incorporate multiple distinct symptoms into one term, inadvertently obscuring patient-level symptoms. A symptom-based approach emphasizes a focus on the patient experience and may better reflect the heterogeneity of the post-acute COVID-19 period.

### Limitations

The symptom lexicon was developed using data from a multi-institution, U.S.-based health care system using a single EHR. Documentation patterns, preferred terminology, and local dialect may differ by geographic location, health system, and EHR vendor. This might impact symptom prevalence or lead to missed symptoms common in other populations. However, the large cohort sample size, and similarities of our findings to those from meta-analyses of prospective and retrospective studies in clinically and geographically distinct populations, supports the external validity and generalizability of this work. Future studies might be needed to replicate the symptom lexicon development pipeline in other EHR systems. Lexicon development was further enriched by manual review to minimize false positive and false negative symptom terminology in the EHR context and to enhance clinical relevance by term consolidation.

Several limitations are inherent in use of EHR data to study post-acute COVID-19 symptoms. First, our study is limited to patients with clinical follow-up in the healthcare system. Patients may have sought out-of-system care post-COVID-19 infection; these patients’ symptoms would not be captured in our lexicon. However, travel advisories during COVID-19 may have limited the ability of patients to seek out of system (or out of state) care, improving our ability to capture encounters in the post-COVID-19 period. Second, the EHR reflects routine care, and therefore, symptoms documented in the clinical notes might be due other visit reasons including underlying diseases (e.g., cancer) or acute events (e.g., stroke) that may or may not be considered sequelae of COVID-19 infection. Although we limited the timeframe of the post-acute COVID-19 phase to day 110, rather than longer timeframes used in other studies (e.g., 6 months) which might otherwise have increased the probability of non-COVID 19 symptom capture, this remains a limitation. Future studies may use computational or epidemiological approaches to investigate correlations between COVID-19 and resulting symptoms.^11^

As detailed in the lexicon development methods, we used keyword search, rather than systematic manual review, for concepts occurring in fewer than 50 patients (0.2% of the population with notes). While rare (low frequency) symptoms may have been missed, these are less likely to be clinically relevant from a population health standpoint.

We also presented limitations of a rule-based NLP approach particularly in consideration of the symptoms’ semantic context. For example, while the NLP tool can accurately identify the symptom term “weight loss” in clinical notes, on chart review we found that the term “weight loss” was often used in the context of weight gain in the post-COVID-19 period, as patients had gained weight and were trialing weight management interventions to decrease their weight. Future studies using word or sentence embeddings in advanced machine learning models may help to extract mentions of symptoms from free-text notes with greater accuracy.^35^ Our proposed symptom lexicon and NLP approach can serve as a base for those future efforts.

## CONCLUSION

We developed a comprehensive post-acute COVID-19 symptom lexicon for EHR data and assessed a lexicon-based NLP approach to extract post-acute COVID-19 symptoms from clinical notes. Further studies are warranted to characterize the prevalence of and risk factors for post-acute COVID-19 symptoms in specific patient populations.

## Supporting information

Supplemental eTable 1

## Data Availability

LW has full access to all of the data in the study and takes responsibility of the integrity of the data and the accuracy of the data analysis. The symptom lexicon from this research work is available through GitHub.

https://github.com/bylinn/Post_Acute_COVID19_Symptom_Lexicon

## FUNDING

No specific funding was received for this project. LW, EM, YCL, DWB, LZ was supported by grant NIH-NIAID R01AI150295 and AHRQ R01HS025375. DF was supported by research funding from IBM Watson (PI: Bates and Zhou) and CRICO.

## AUTHOR CONTRIBUTIONS

LW has full access to all of the data in the study and takes responsibility of the integrity of the data and the accuracy of the data analysis. *Concept and design*: LW, DF. *Acquisition, analysis, or interpretation of data*: All authors. *Draft of the manuscript*: LW, DF. *Critical revision of the manuscript for important intellectual content*: LW, DF, DWB, LZ. *Statistical Analysis*: LW, DF. *Obtained funding*: Not applicable. *Administrative, technical, or material support*: LZ. S*upervision*: DWB, LZ.

## CONFLICT OF INTEREST STATEMENT

LW, DF, EM, YCL, LZ reports no disclosures. DWB reports grants and personal fees from EarlySense, personal fees from CDI Negev, equity from ValeraHealth, equity from Clew, equity from MDClone, personal fees and equity from AESOP, personal fees and equity from Feelbetter, and grants from IBM Watson Health, outside the submitted work.

## ADDITIONAL INFORMATION

The post-acute COVID-19 symptom lexicon can be accessed at: https://github.com/bylinn/Post_Acute_COVID19_Symptom_Lexicon.

## Notes

### Author Declarations

This study was reviewed by the Mass General Brigham Human Research Committee and determined to be exempt/non-human subjects research (Protocol 2020P000816).

