## Supplemental eTable 1 for "Development of a Post-Acute Sequelae of COVID-19 (PASC) Symptom Lexicon Using Electronic Health Record Clinical Notes"

**Supplemental Material Contents:**

**eTable 1.** Challenges in a rule-based natural language processing (NLP) approach to symptom extraction from electronic health record (EHR) clinical notes

**eTable 1.** Challenges in a rule-based natural language processing (NLP) approach to symptom extraction from electronic health record (EHR) clinical notes

| EHR Feature | Example from clinical notes | Challenge for NLP |
|---|---|---|
| Symptoms scores (Patient Reported Outcome Measures) | “Edinburgh Depression Scale score:2” | Requires medical knowledge to determine if the patient is having depression symptoms |
| Standard instructions | “If you find you often feel sad or hopeless, talk with your doctor”<br><br>“Symptoms of depression include: a sad or gloomy mood, lack of pleasure in activities you previously enjoyed, difficulties managing your weight, early morning awakening with inability to return to sleep or excessive sleeping, agitation or moving so slow that others notice, difficulty concentrating, fatigue, or mental slowness” | Often included in clinical notes as anticipatory guidance to patients based on reason for visit or medication prescriptions, regardless of current patient symptoms |
| Linguistic context | “Stress test”<br><br>“Stress fracture”<br><br>“The patient stressed that the nausea had not gone away despite dietary changes” | Symptom terminology may also be used in other medical contexts or as colloquialisms |
| Negation | “Mental Health Screening: Depression Concerns: none”<br><br>“Fevers / Chills: n”<br><br>“(-) weight gain” | Negation terms may be distant from symptom term, or may be represented by abbreviations [n; (-)] |
| Past medical history | “DOE (dyspnea on exertion) 9/9/2019” | Symptoms preceded the study timeframe |
| Treatment indication | “albuterol 90 mcg/actuation inhaler Inhale 2 puffs into the lungs every 6 (six) hours as needed for wheezing or shortness of breath/dyspnea” | Medication instruction include clinical indications, which may be representative of active or anticipated patient symptoms |
| Allergic reaction | “Statins-hmg-coa reductase inhibitors Myalgia 10/06/2020” | Symptoms in the study timeframe but with a |

| | | |
|---|---|---|
| | | clear non-COVID-19 associated trigger |
| Review of systems | "[] Anorexia [] fatigue [x] insomnia [] fever [] chills [] dizziness [] weight gain [x] weight loss [] night sweats" | Clinical tabulation of current symptoms denoted with signs such as "+" or "x" |
| Implied symptoms | "Encouraged to lose weight, portion control, hydration, and exercise will be key"<br><br>"She has intentionally lost weight, reports an 8lb weight loss with cutting out soda and walking more" | Therapeutic intervention implies presence of a symptom, e.g., a patient needs to lose weight, because they have had weight gain. Rule based NLP may have difficulty differentiating intentional and unintentional patient reported outcomes or symptoms |
